# A Novel Temperature Controlled Bipolar Radiofrequency Ablation: An Ex Vivo Study for Optimizing Efficacy and Safety Parameters

**DOI:** 10.1101/2024.11.11.24317133

**Authors:** Osamu Inaba, Yukihiro Inamura, Takamitsu Takagi, Shin Meguro, Kentaro Nakata, Toshiki Michishita, Yuhei Isonaga, Shinichi Tachibana, Hiroaki Ohya, Akira Sato, Shinsuke Miyazaki, Yasuteru Yamauchi, Masahiko Goya, Junichi Nitta, Tetsuo Sasano

## Abstract

**Background:** Bipolar radiofrequency catheter ablation (BRFA) is a potential treatment for refractory ventricular arrhythmias from deep myocardial tissue. However, clear indicators of efficacy and safety remain undefined.

**Methods:** In an ex vivo model, BRFA was performed using either the QDOT Micro™ (QDT) or Thermocool SmartTouch SF™ as the active catheter, with the DiamondTemp Ablation™ (DTA) as the return catheter. Predictors of transmural lesion formation and steam-pop occurrence were assessed.

**Results:** A total of 391 BRFA applications were performed with variations in the interelectrode distance between the active and return catheter tips, ranging from 6 to 27 mm, under various catheter tip and tissue contact configurations. The ablation index (AI) adjusted for inter-electrode distance effectively predicted transmural lesion formation. Logistic regression analysis revealed a coefficient for AI of −0.040 (SE: 0.0067; 95% CI: [−0.053, −0.027]; p < 0.0001) and for inter-electrode distance of 2.2 (SE: 0.35; 95% CI: [1.5, 2.9]; p < 0.0001). The decision boundary for transmural lesion formation was AI = 54 × inter-electrode distance – 260. When AI exceeded this value, sensitivity, specificity, and positive and negative predictive values for predicting transmural lesions were 89%, 92%, 91%, and 90%, respectively. When the AI was further increased by 50, the specificity reached 100%.

Steam-pops on the active catheter side occurred only during power-controlled BRFA and were absent in temperature-controlled BRFA with a 45°C cutoff. On the return side, steam-pops occurred when DTA temperature exceeded 55°C, with deeper cracks observed above 60°C.

**Conclusions:** An AI adjusted for inter-electrode distance strongly predicted transmural lesions. Temperature-controlled BRFA with a 45°C cutoff for QDT as the active catheter and 55°C for DTA as the return catheter may prevent steam-pops. Additionally, steam-pops occurring at higher catheter tip temperatures were associated with deeper tissue cracks.

**Clinical Perspective:** *What is known?*

- Bipolar radiofrequency ablation has shown promise as an effective treatment option for ventricular arrhythmias originating from deep regions of the ventricular myocardium.
- The indicators for successful transmural lesion formation and the predictors for steam-pop are not yet well understood, and the accuracy of conventional metrics remains limited.

*What the study adds*

- The ablation index (AI) necessary for achieving successful transmural lesion formation during bipolar radiofrequency ablation, using a QDOT micro™ (QDT) as the active catheter and a DiamondTemp Ablation™ (DTA) the return catheter, can be determined using the formula: AI = 54 × inter-electrode distance – 260, where the inter-electrode distance denotes the distance between the active and return electrodes.
- During bipolar radiofrequency ablation, performing temperature-controlled ablation with a temperature cutoff of 45°C for the QDT and 55°C for the DTA may help prevent steam-pop.
- Furthermore, the higher the DTA temperature at the time of steam-pop, the deeper the cracks that are formed.

## Introduction

Radiofrequency (RF) catheter ablation is currently considered an effective option for the treatment of ventricular arrhythmias (VAs). Conventional unipolar radiofrequency ablation, which can be performed in conjunction with a variety of catheters and 3D mapping systems, is the standard method. However, its efficacy is limited when the origin of the VAs is intramural.

Bipolar RF catheter ablation (BRFA) is one of the next options for applying radiofrequency current to areas where it is difficult to form lesions deep enough using conventional unipolar ablation. (1) In BRFA, a RF current is passed between two catheters, an active catheter connected to the active port of the RF generator, and a return catheter connected to the indifferent electrode port instead of a return electrode patch, making it possible to form a deep ablation lesion between the two electrodes. The efficacy and safety of BRFA is gradually being recognized (2–5), but the cables connecting the generator and the return electrode are not widely commercially available and often have to be custom made. Furthermore, there are currently no established ablation settings or safety indicators for BRFA, and, because this is off-label use, catheter manufacturers also do not have sufficient data. Because serious complications of BRFA have been reported, it is important to establish efficacy and safety parameters of BRFA (6,7).

For conventional unipolar RF ablation, the ablation index (AI) was developed as a quantitative ablation lesion predictor, that includes contact force (CF), RF application time, and RF power in a weighted formula. The first aim of this study examined whether AI could be used to predict intermural lesion formation in BRFA as an efficacy parameter.

For a safety indicator, we focused on monitoring electrode temperature during BRFA. Recently, we reported a case of ventricular tachycardia in which temperature-controlled BRFA was successfully performed from both the endocardium and epicardium using a diamond embedded tip catheter (DiamondTemp Ablation [DTA] system [Medtronic, Inc., Minneapolis, Minnesota, USA]). (8) In temperature-controlled mode, the RF output is attenuated as the tissue surface temperature increases, potentially preventing steam-pop. DTA is open-irrigated but can measure the temperature of the tissue surface with high sensitivity, making temperature-controlled ablation possible.

Furthermore, in recent years, QDOT micro^TM^ (QDT); Biosense Webstar, Irvine, California, USA have become widely available, enabling temperature-controlled ablation using the CARTO (Biosense Webstar, Irvine, California, USA). In the latter aim of the study, the safety of temperature-controlled BRFA and the relationship between electrode temperature and steam-pop during BRFA were investigated.

## Methods

Fresh porcine ventricles were obtained commercially within 48 hours of sacrifice. A porcine ventricular tissue was fixed in a water pool filled with 37-degree saline, and an active catheter was perpendicularly placed on the endocardial side and a return catheter on the epicardial side. In this ex vivo study, we used a Thermocool SMARTTOUCH SF^TM^ (STSF); Biosense Webstar, Irvine, California, USA or a QDT, which is capable of monitoring AI during RF application as the active catheter and a DTA as the return catheter to examine the usefulness of the ablation index in BRFA using the CARTO system.

An active catheter was connected to the active port of the radiofrequency generator of CARTO (nGEN, Biosense Webstar, Irvine, California, USA), and DTA as a return catheter was connected to ground electrode port of nGEN via generator connection box and pin box using alligator clip. (Figure 1 Main) This study was divided into two parts. In the first part, to investigate the optimal AI value for forming an intermural lesion, QDT and DTA were attached to ventricular muscle tissue in four configurations: both perpendicular with slight angle, QDT perpendicular and DTA parallel, both parallel, and QDT parallel and DTA perpendicular (Model 1A, B, C, D, Figure 1 A, B, C, D). In this process, since previous studies have reported that proper attachment of the second ring electrode in DTA is crucial, it was slightly angled to ensure the second electrode contacted with the tissue before applying the BRFA when attaching DTA perpendicularly. (9)

**Figure 1.**
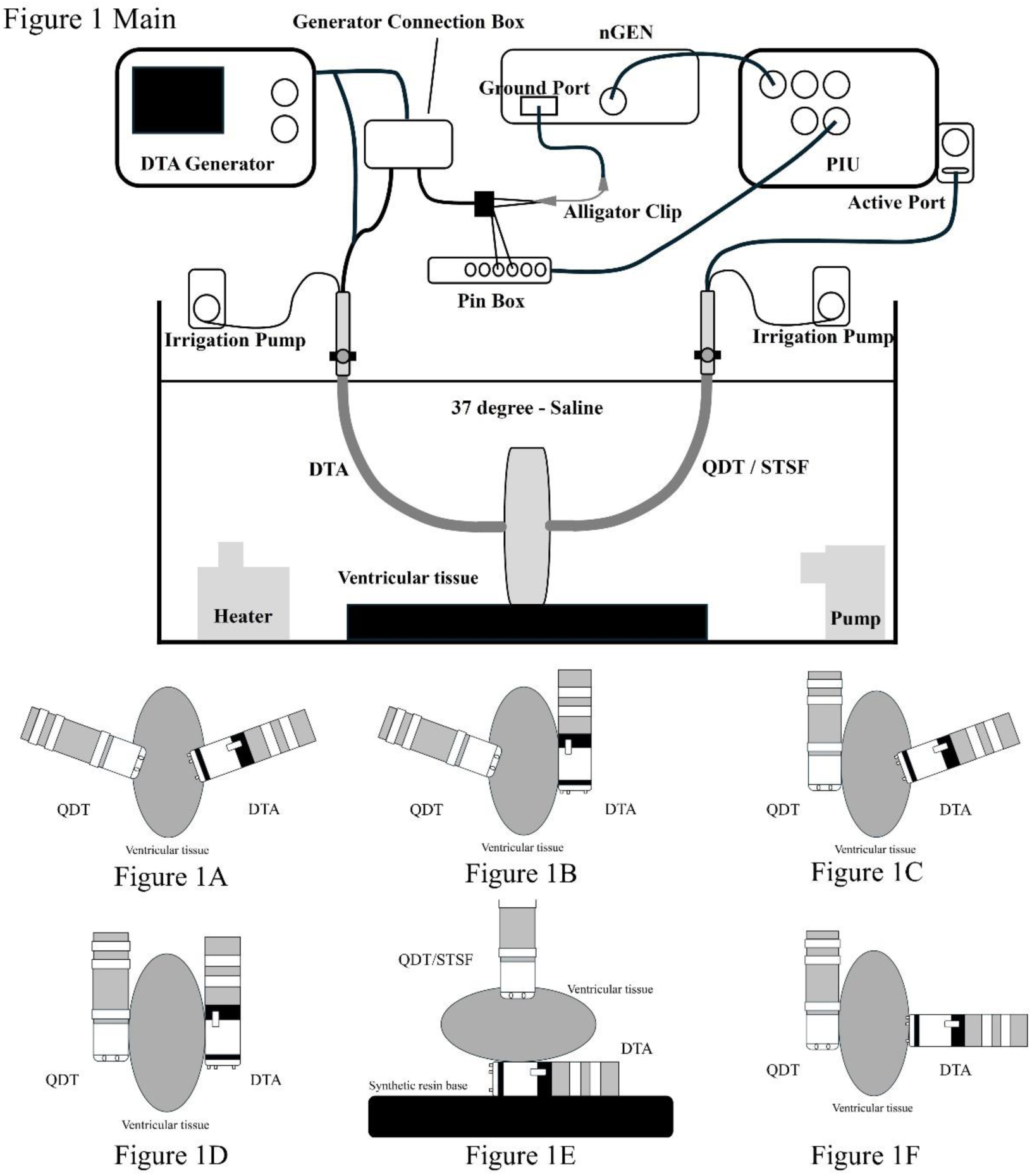
The circuit model for BRFA using CARTO and DTA is shown. The QDOT/STSF is connected as the active catheter, attached to the active port in the usual manner. The DTA is connected to the ground port via an alligator clip. To display the DTA on the CARTO system during the experiment, only the proximal electrodes of the DTA are connected to the patient interface unit of the CARTO system via the Pin box. Furthermore, the contact patterns between each catheter tip and the ventricular myocardium are shown in Figures 1A through 1F. Figures 1A to 1D illustrate four different configurations where the QDOT and DTA were adhered perpendicularly and parallel (Models 1A - 1D). Figure 1E shows a scenario in which the DTA was sandwiched between the ventricular myocardium and a synthetic resin base to create conditions prone to steam-pop formation (Model 2). Figure 1F shows a special configuration in which only the 0.6 mm long distal small tip of the DTA is in contact with the ventricular myocardium, while the proximal large tip is not in contact with the tissue (Model 3). DTA, DiamondTemp Ablation^TM^; nGEN, nGEN^TM^ generator; PIU, patient interface unit; QDT. QDOT micro^TM^; STSF, Thermocool SMARTTOUCH SF^TM^.

The purpose of the second part was to examine steam-pop prediction. Both QDT and STSF were used as active catheters, and to make audible steam-pop more likely to occur, the DTA electrode was inserted between the myocardium and a synthetic resin base and attached parallel to the myocardium to concentrate the RF current in a narrower area and make it easier to raise the tissue temperature. (Model 2, Figure 1E).

In addition to these ablation sets, another lesion set was examined where only the distal thin-tip electrode of the DTA was completely perpendicularly attached to the tissue for current application. (Model 3, Figure 1F)

In each model, the target AI was set, and ablation was started with a CF of about 5 to 15 g and a power output of 25 to 40 W. For all ablations using QDT, QMODE^TM^ was used with a temperature cutoff set at 45 °C. The irrigation flow for BRFA using the QDT was regulated by temperature control with QMODE^TM^, while for BRFA using the STSF, it was set to 15 ml/min higher than 30W, and 8 ml/min at 30W or lower. The irrigation flow for the DTA was manually set to 8 ml/min during RF delivery. Ablation was stopped when an audible steam-pop occurred. The temperature and impedance of the DTA were visually monitored and recorded.

After ablation, myocardial sections were cut and visually inspected to confirm whether transmural lesions were formed. Additionally, the distance between the two electrodes where BRFA was performed was measured. The target AI was gradually increased from a lower initial value until transmural lesions were formed. For sections where a steam-pop occurred, the specific side of the catheter where the steam-pop occurred was visually recorded in addition to noting the tactile sensation experienced by the catheter operator. Furthermore, the extent of the cracks was also measured. During lesion confirmation, the measurers were blinded to the AI and the type of active catheter used.

In the examination of the effectiveness of AI, for both Model 1 and Model 2, the AI, the distance between electrodes of QDT/STSF and DTA used in BRFA (inter-electrode distance), CF, BRFA duration, energy, circuit impedance drop rate (% Impedance drop), temperature of the DTA and the values of AI, BRFA duration, and energy divided by the inter-electrode distance (AI/inter-electrode distance, BRFA duration/inter-electrode distance and energy/inter-electrode distance) were investigated for ablations that resulted in transmural lesions and those that did not.

In the examination of steam-pops, the same parameters were investigated for sections where steam-pops occurred and those where they did not. Additionally, the depth of the cracks caused by steam-pop was classified into four categories: less than 25% of the surface or not visible to the naked eye, 25% to 50% of the surface, more than 50% of the surface with partial incompleteness, and through-wall cracks. This study was performed in accordance with the Guide for the Care and Use of Laboratory Animals published by the U.S. National Institutes of Health.

### Statistical Analysis

Continuous data are given as mean ± standard deviation if normally distributed or the median, maximum, and minimum values if not normally distributed. Categorical data are given as counts and percentages. Continuous data were compared using Student’s t test or one-way analysis of variance when normally distributed or Mann-Whitney U test when not normally distributed. Categorical data were compared using χ^2^ test across the groups.

Factors related to the formation of transmural lesions were analyzed using univariate analysis, and the relationship between those factors and inter-electrode distance was examined on a scatter plot. Furthermore, on that scatter plot, the BRFA application groups that could and could not form transmural lesions were depicted separately. A logistic regression model was employed to determine the decision boundary between the groups. The decision boundary, where the probability of an observed value belonging to the transmural lesion group is 0.5, was plotted, and the equations related to the formation of transmural lesions based on the inter-electrode distance required for lesion formation were derived. Additionally, the sensitivity, specificity, and positive and negative predictive values of BRFA applications beyond the AI threshold—calculated based on electrode distance using the decision boundary formula—were assessed for their effectiveness in predicting transmural lesion formation. The level of significance was set at P<0.05.

Analyses were performed with JMP^®^ 14 (SAS Institute Inc., Cary, NC).

## Results

### Evaluation of AI and Transmural Lesion Formation

In total, 391 BRFA were performed on slices of varying thickness with inter-electrode distances ranging from 6 to 27 mm, and no damage occurred to the CARTO system or the generators of CARTO and DTA. Two-hundred-thirty-three ablations were performed in Model 1 and 148 ablations in Model 2, resulting in 127 transmural lesions in Model 1 and 109 transmural lesions in Model 2.

In Model 1 overall, except for the average CF, average power and maximum temperature of DTA, all parameters showed higher values in the ablations where transmural lesions were formed (AI: 680±240, 580±200, p=0.0005; AI/inter-electrode distance: 46±7.6, 32±5.8, p<0.0001; BRFA duration: 64 [6.2 - 630], 40 [3.7 - 520], p=0.0004; BRFA duration/inter-electrode distance: 4.7 [0.62 - 23], 2.2 [0.32 - 21], p<0.0001; Energy: 2300 [220 - 220000], 1500 [130 - 14000], p=0.0011; Energy/inter-electrode distance: 150 [11 - 820], 100 [12 - 560], p<0.0001; maximum % Impedance drop: 16±4.1, 13±3.4, p<0.0001; inter-electrode distance: 14 [6 - 27], 18 [8 - 27], p<0.0001) (Table 1). Among these, only AI/interelectrode distance and BRFA duration/inter-electrode distance showed significantly higher values in the ablations where transmural lesions were formed across all variations of catheter tip configurations from Model 1A to 1D. (Supplemental Table 1)

**Table 1.**
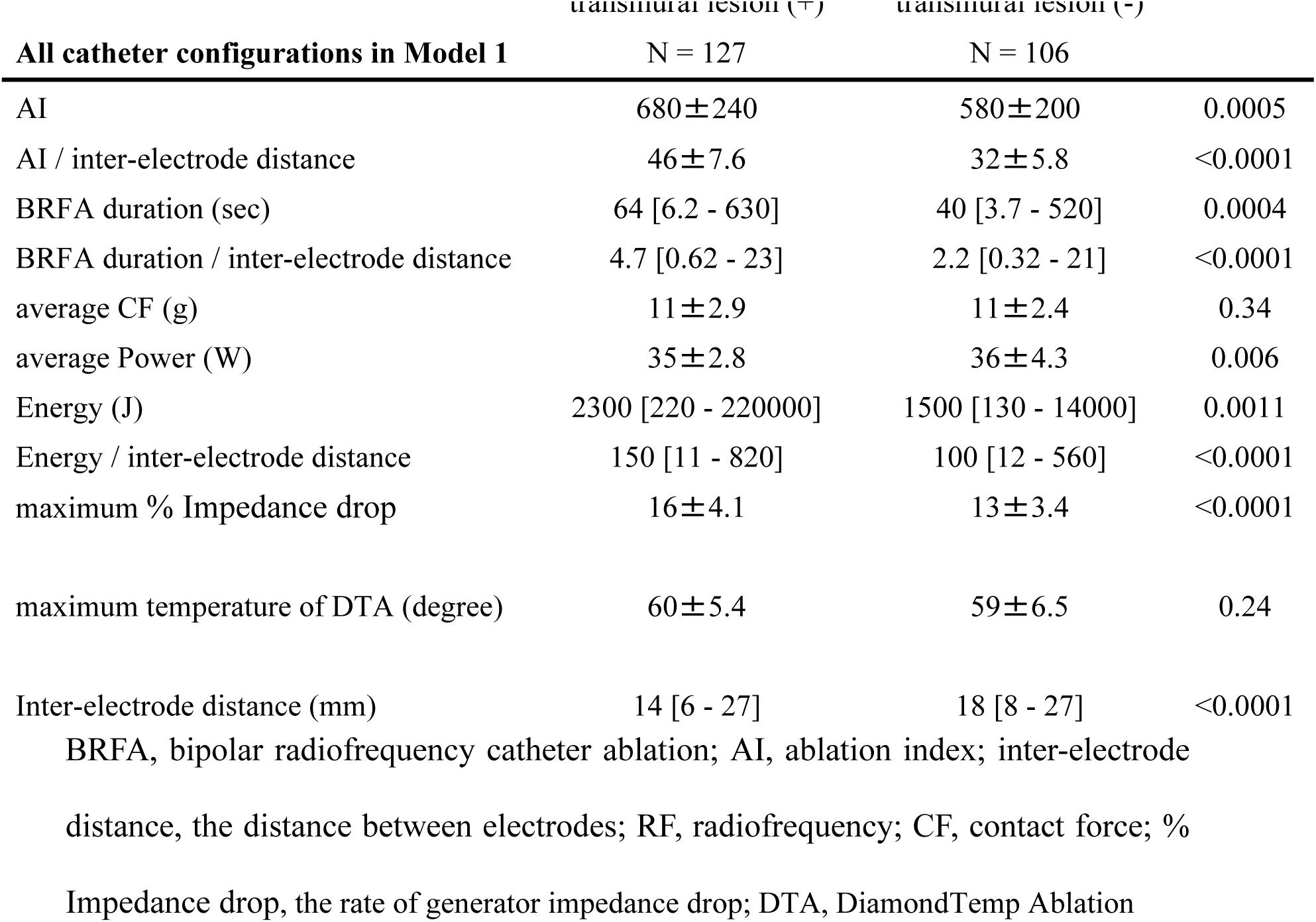
Comparison of BRFA parameters between ablations with and without transmural lesion formation.

|  | transmural lesion (+) | transmural lesion (-) |  |
| --- | --- | --- | --- |
| **All catheter configurations in Model 1** | **N = 127** | **N = 106** |  |
| AI | 680±240 | 580±200 | 0.0005 |
| AI / inter-electrode distance | 46±7.6 | 32±5.8 | <0.0001 |
| BRFA duration (sec) | 64 [6.2 - 630] | 40 [3.7 - 520] | 0.0004 |
| BRFA duration / inter-electrode distance | 4.7 [0.62 - 23] | 2.2 [0.32 - 21] | <0.0001 |
| average CF (g) | 11±2.9 | 11±2.4 | 0.34 |
| average Power (W) | 35±2.8 | 36±4.3 | 0.006 |
| Energy (J) | 2300 [220 - 220000] | 1500 [130 - 14000] | 0.0011 |
| Energy / inter-electrode distance | 150 [11 - 820] | 100 [12 - 560] | <0.0001 |
| maximum % Impedance drop | 16±4.1 | 13±3.4 | <0.0001 |
| maximum temperature of DTA (degree) | 60±5.4 | 59±6.5 | 0.24 |
| Inter-electrode distance (mm) | 14 [6 - 27] | 18 [8 - 27] | <0.0001 |

BRFA, bipolar radiofrequency catheter ablation; AI, ablation index; inter-electrode distance, the distance between electrodes; RF, radiofrequency; CF, contact force; % Impedance drop, the rate of generator impedance drop; DTA, DiamondTemp Ablation

As the inter-electrode distance increases, the AI required for transmural lesion formation increases linearly. (Figure 2A) Logistic regression analysis revealed that the regression coefficient for AI was −0.040 (standard error: 0.0067; 95% confidence interval: [−0.053, −0.027]; p < 0.0001), while the regression coefficient for inter-electrode distance was 2.2 (standard error: 0.35; 95% confidence interval: [1.5, 2.9]; p < 0.0001). Based on these results, the decision boundary for achieving or not achieving transmural lesion was calculated as: AI = 54 × inter-electrode distance − 260. Out of the 127 ablations where transmural lesions were formed, 117 ablations had AI values exceeding the decision boundary formula, while out of the 106 ablations where transmural lesions were not formed, 94 ablations had AI values below the decision boundary formula. The sensitivity, specificity, and positive and negative predictive values of the decision boundary for transmural lesion formation were 89%, 92%, 91%, and 90%, respectively. The relationship between AI and inter-electrode distance for ablations that achieved transmural lesion formation and those that did not, in each catheter tip configuration, is shown in the Supplemental Figure 1.

**Figure 2.**
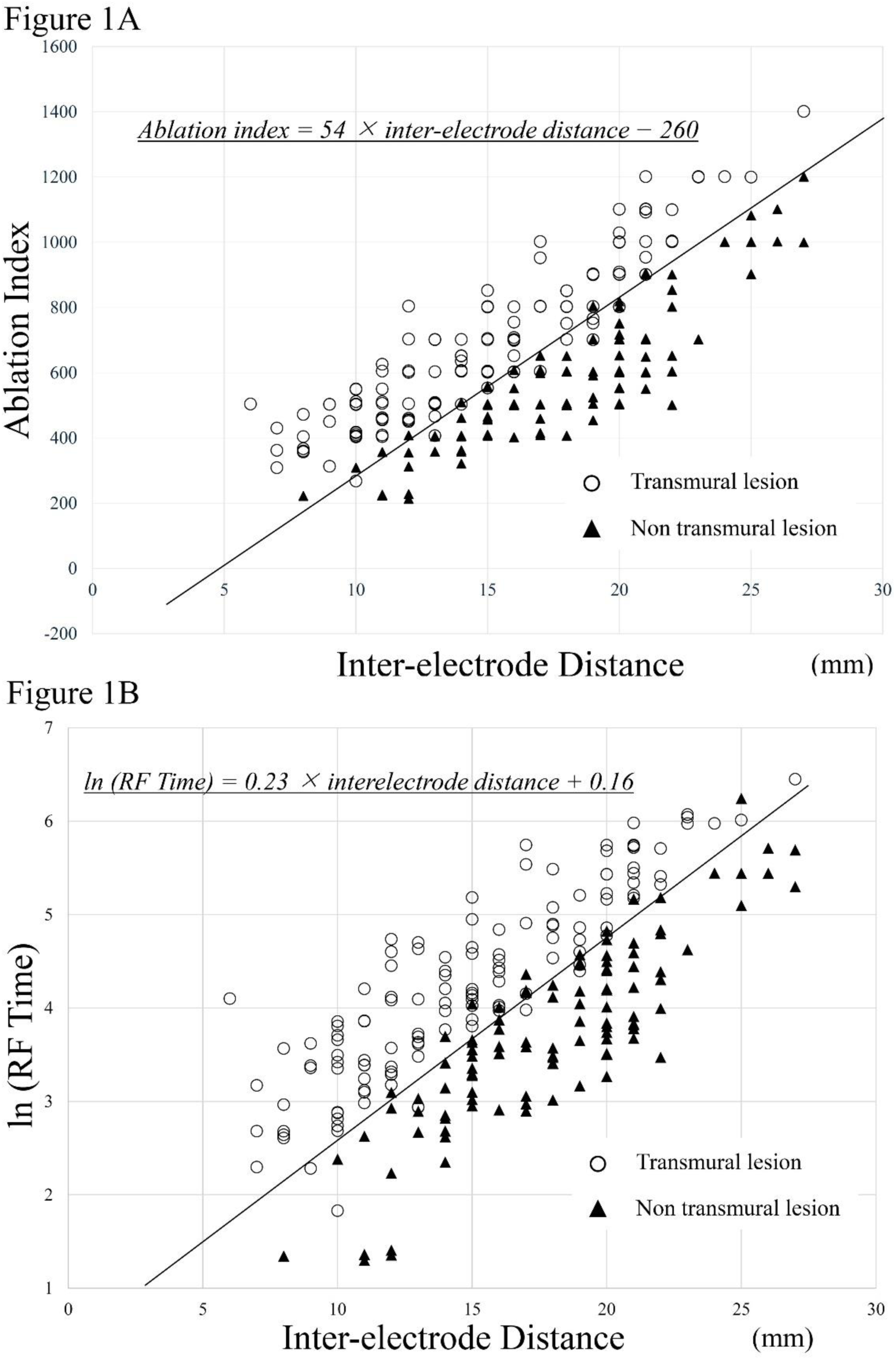
The relationship between inter-electrode distance, AI, and the logarithm of BRFA duration is displayed, separated into ablations with and without transmural lesion formation. The AI and ln (BRFA duration) required to achieve transmural lesion formation increase linearly as the inter-electrode distance increases. The decision boundary equations for achieving and not achieving transmural lesion formation are AI = 54 × inter-electrode distance − 260 and ln (BRFA duration) = 0.23 × interelectrode distance + 0.16, respectively. AI, ablation index; BRFA, bipolar radiofrequency catheter ablation; inter-electrode distance, the distance between electrodes used in BRFA.

Regarding the analysis of BRFA duration, it was observed from the scatter plot that taking the natural logarithm of BRFA duration (ln (BRFA duration)) reveals a linear boundary between ablations that formed transmural lesions and those that did not. In logistic regression analysis, the regression coefficient for the natural logarithm of BRFA duration was −7.0 (standard error: 1.1; 95% confidence interval [−9.3, −5.1], p < 0.0001), and the regression coefficient for the inter-electrode distance was 1.6 (standard error: 0.25; 95% confidence interval [1.2, 2.2], p < 0.0001). The decision boundary for achieving or not achieving transmural lesion was calculated as: ln (BRFA duration) = 0.23 × inter-electrode distance + 0.16. (Figure 2B) Similarly to the AI analysis, the sensitivity, specificity, and positive and negative predictive values for the decision boundary for transmural lesion formation based on ln (BRFA duration) were calculated as 95%, 85%, 88%, and 94%, respectively.

### Evaluation of Steam-Pop

In Model 2, A total of 123 temperature-controlled ablations using QDT and 25 power-controlled ablations using STSF were performed, and 46 steam-pops were observed. Meanwhile, in Model 1, where all ablations were performed with temperature control, only 3 steam-pops occurred out of 233 applications.

All the steam-pops in Model 1 occurred on the return catheter side 47 to 67 seconds after the start of the ablation. At that time, the % Impedance drop was 17 to 21%, the temperature of DTA was 58 to 71 degrees, and the crack penetration depth was less than 50%. In Model 2, a comparison between 102 ablations without steam-pop and 46 ablations with steam-pop revealed that temperature-controlled ablations were more frequent in the ablations without steam-pop, whereas power-controlled ablations were more common in the ablation with steam-pop. In temperature-controlled ablations, all 32 steam-pops occurred on the return electrode side, while in power-controlled ablations, one steam-pop occurred on the active electrode side and the last 13 steam-pops occurred on the return electrode side. The ablations with steam-pops had significantly higher for average power, % Impedance drop, and DTA temperature compared to the ablations without steam-pops (Average Power: 28±10, 35±5.9, p<0.0001; maximum % Impedance drop: 22±6.2, 25±3.7, p=0.0007; maximum temperature of DTA: 60±7.7, 66±5.3, p<0.0001) (Table 2).

**Table 2.**
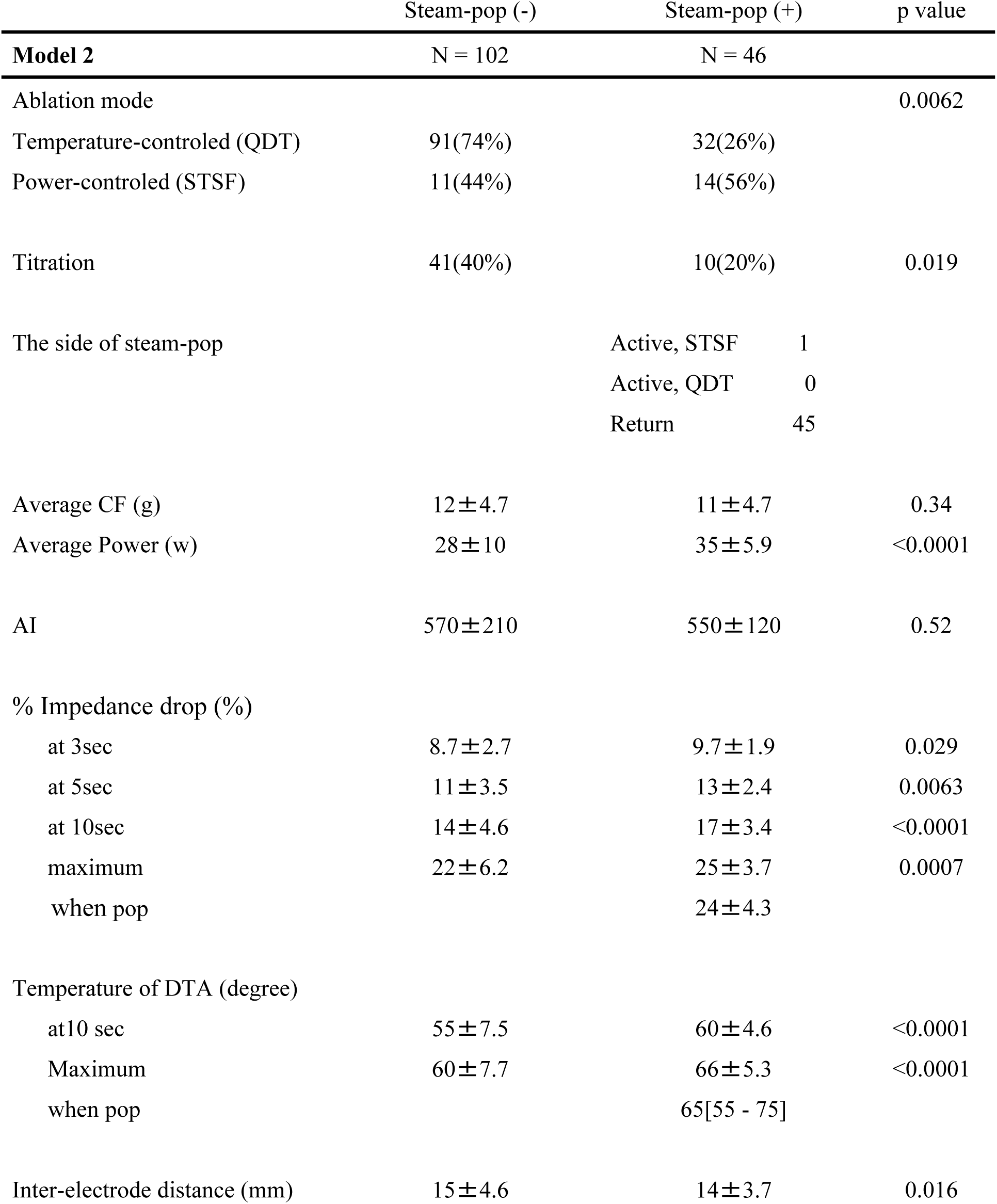
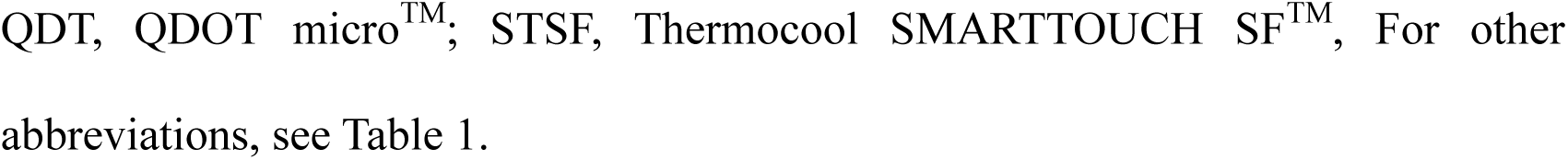
Comparison of protocols and parameters between ablations with and without steam-pop occurrence in Model 2.

|  | Steam-pop (-) | Steam-pop (+) | p value |
| --- | --- | --- | --- |
| **Model 2** | N = 102 | N = 46 |  |
| Ablation mode |  |  | 0.0062 |
| Temperature-controlled (QDT) | 91(74%) | 32(26%) |  |
| Power-controlled (STSF) | 11(44%) | 14(56%) |  |
| Titration | 41(40%) | 10(20%) | 0.019 |
| The side of steam-pop |  | Active, STSF 1<br>Active, QDT 0<br>Return 45 |  |
| Average CF (g) | 12±4.7 | 11±4.7 | 0.34 |
| Average Power (w) | 28±10 | 35±5.9 | <0.0001 |
| AI | 570±210 | 550±120 | 0.52 |
| % Impedance drop (%) |  |  |  |
| at 3sec | 8.7±2.7 | 9.7±1.9 | 0.029 |
| at 5sec | 11±3.5 | 13±2.4 | 0.0063 |
| at 10sec | 14±4.6 | 17±3.4 | <0.0001 |
| maximum | 22±6.2 | 25±3.7 | 0.0007 |
| when pop |  | 24±4.3 |  |
| Temperature of DTA (degree) |  |  |  |
| at10 sec | 55±7.5 | 60±4.6 | <0.0001 |
| Maximum | 60±7.7 | 66±5.3 | <0.0001 |
| when pop |  | 65[55 - 75] |  |
| Inter-electrode distance (mm) | 15±4.6 | 14±3.7 | 0.016 |

QDT, QDOT micro<sup>TM</sup>; STSF, Thermocool SMARTTOUCH SF<sup>TM</sup>, For other abbreviations, see Table 1.

Among the 46 steam-pops that occurred in Model 2, an analysis of the 45 instances that occurred on the DTA side revealed that all steam-pops occurred when the DTA temperature 55 °C or higher, and the % Impedance drop exceeded 13Ω. Steam-pop did not occur when the DTA temperature was below 55°C or when the % Impedance drop was less than 13Ω. Figure 3A shows a plot of the maximum temperature of the DTA during ablation where steam-pop did not occur and the DTA temperature at the time of steam-pop during ablation where steam-pop occurred. When the DTA temperature cutoff is set at 55°C, the sensitivity, specificity, positive predictive value, and negative predictive value for predicting steam-pop occurrence are 100%, 27%, 38%, and 100%, respectively.

**Figure 3.**
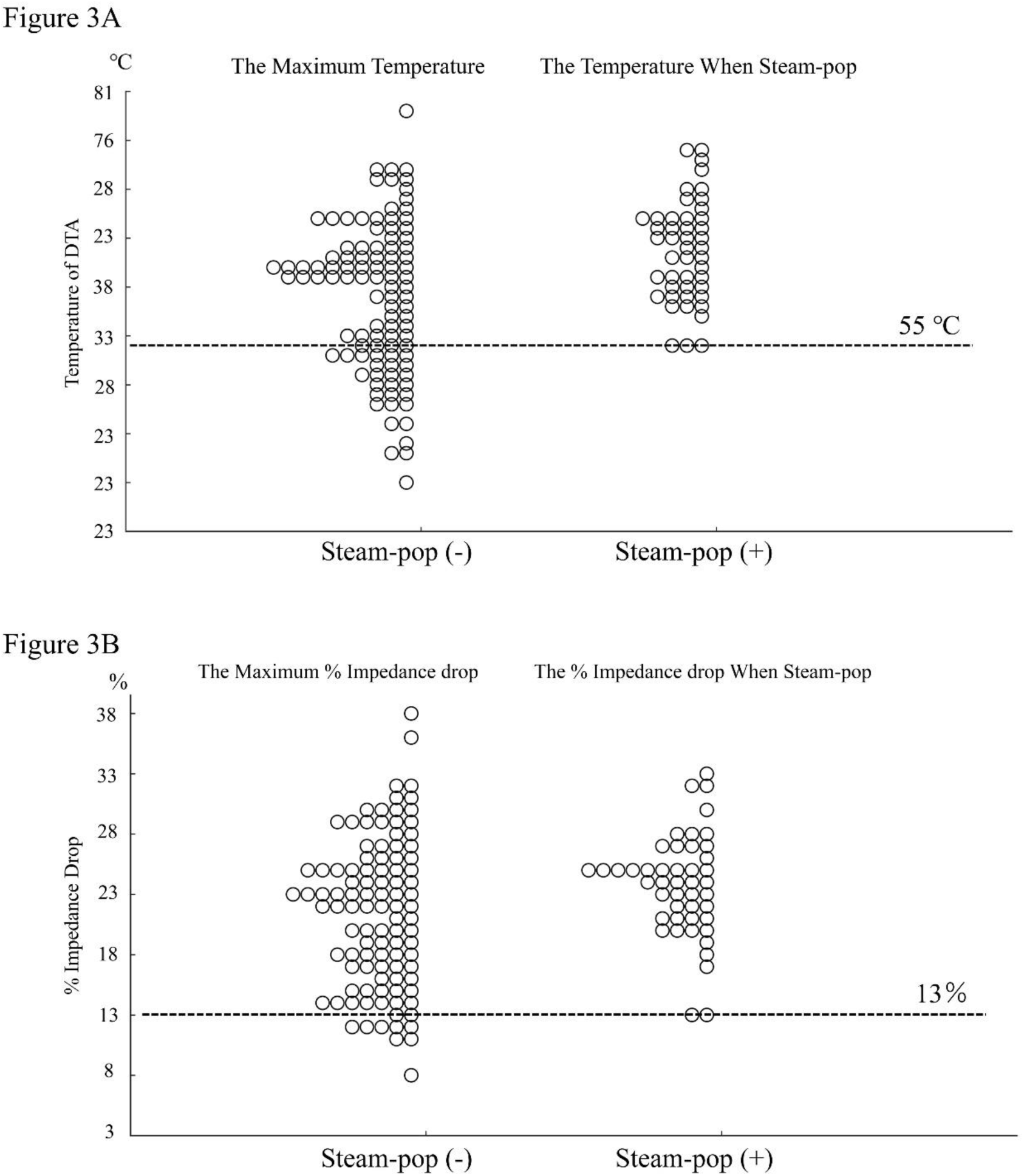
Comparison of the maximum DTA temperature and maximum % Impedance drop in ablation without steam-pop, and the DTA temperature and % Impedance drop at the time of steam-pop in ablation where steam-pop occurred. Figure 3A shows that all steam-pops occurred only when the DTA temperature was 55°C or higher. Similarly, Figure 3B shows that all steam-pops occurred only when the % Impedance drop was 13% or higher. DTA, DiamondTemp Ablation^TM^; %Impedance drop, the rate of generator impedance drop.

Similarly, Figure 3B shows a plot of the % Impedance drop. When the %Imp drop cutoff is set at 13%, the sensitivity, specificity, positive predictive value, and negative predictive value for predicting steam-pop occurrence are 100%, 8.8%, 33%, and 100%, respectively.

Furthermore, the higher the DTA temperature when steam-pop occurred, the larger the cracks that formed. The cracks penetrating 25% or more of the depth were observed only when the DTA temperature was 60 °C or higher. Cracks confined to the mid-myocardium were not observed in these ablations. (Figure 4A) A similar result was also observed with the % Impedance drop at steam-pop. Cracks that extended beyond 50% were only formed when the % Impedance drop exceeded 21%. (Figure 4B)

**Figure 4.**
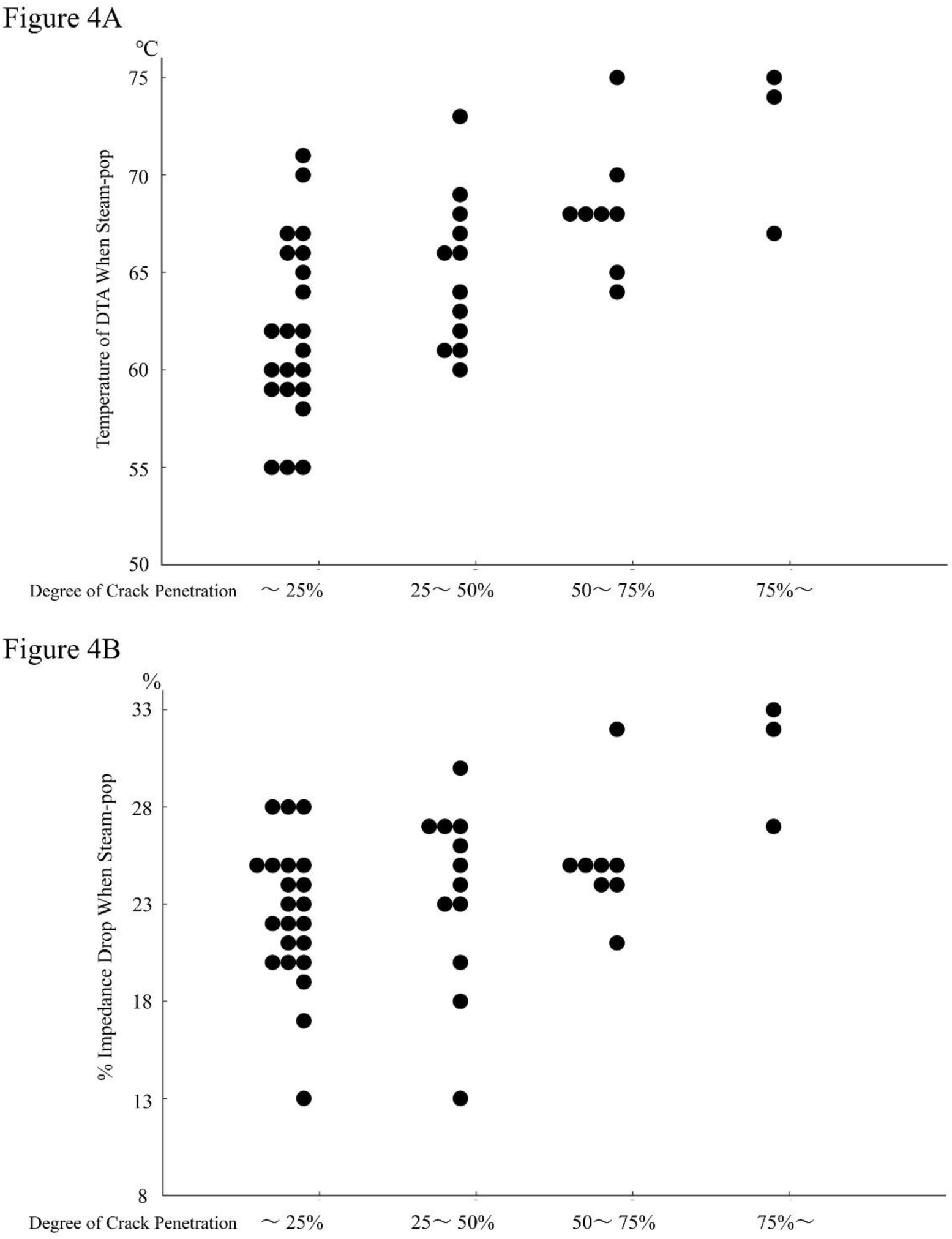
The relationship between the depth of visually confirmed cracks in ablation where steam-pop occurred, and the DTA temperature and % impedance drop at the time of steam-pop. Cracks with a depth of 25% or more were observed only when steam-pop occurred with the DTA temperature reaching 60°C or higher. (Figure 4A) Similarly, cracks with a depth of 50% or more were observed only when the % impedance drop increased by 21% or more. (Figure 4B). Abbreviations are the same as in Figure 3.

### Model 3: A special catheter configuration

Lastly, 10 BRFA procedures were performed in Model 3, where only the distal 0.6 mm electrode of the DTA was attached to the myocardial tissue, and the second electrode was not attached. BRFA procedures with AI 400-750 were applied to myocardial tissue with an inter-electrode distance of 11 mm to 15 mm. However, no transmural lesions were formed, and lesion formation on the return electrode side was either absent or minimal. The highest temperature of the DTA remained below 46 °C. (Table 3)

**Table 3.** Ablation parameters and lesion depth and size on each electrode side during BRFA performed with only the distal small tip of the DTA in contact with the tissue.

| **Model 3** | **N = 10** |
| --- | --- |
| AI | 520±120 |
| AI / inter-electrode distance | 36±8.2 |
| BRFA duration (sec) | 32 [19 - 72] |
| RF time / inter-electrode distance | 2.5 [1.3 - 5.1] |
| average CF (g) | 11±4.1 |
| average Power (W) | 35 |
| Energy (J) | 1100 [670 - 2500] |
| Energy / inter-electrode distance | 87 [45 - 180] |
| maximum % Impedance drop (%) | 8.6±1.6 |
| maximum temperature of DTA (degree) | 45±0.67 |
| Inter-electrode distance (mm) | 15 [11 - 15] |
| Active electrode |  |
| lesion depth (mm) | 3.9±1.0 |
| lesion short axis (mm) | 5.8±1.1 |
| Return electrode |  |
| lesion depth (mm) | 0 [0 - 2] |
| lesion short axis (mm) | 0 [0 - 2] |

Abbreviations are the same as in Table 1 and 2.

## Discussion

This study is the first ex vivo study to investigate transmurality and steam-pop occurrence in BRFA using QDT and DTA. This study has three notable points. First, it was found that performing BRFA using QDT as the active catheter and DTA as the return catheter is likely to not cause malfunctions with the system used in this study. As previously mentioned, BRFA is an off-label use, and the catheter manufacturers do not have data regarding potential system malfunctions. Therefore, the fact that nearly 400 BRFA procedures did not cause any issues with the CARTO system or the DTA generator is valuable information.

Second, this is the first report to confirm that AI can predict transmural lesion formation in BRFA. Previous studies have reported that factors related to transmurality in BRFA include total energy (10), thickness of the myocardium (11), use of a 3.5mm tip irrigated catheter (11), BRFA duration (12), impedance drop (12) and contact surface area of the return catheter (13). The results of the present study indicated that the inter-electrode distance and the corresponding AI or BRFA duration were crucial in transmural lesion formation. As the inter-electrode distance increased, the AI and the ln (BRFA duration) required to achieve transmural lesion formation increased linearly up to an inter-electrode distance of 27 mm. In this study, both AI and ln (BRFA duration) yielded highly accurate predictive formulas for achieving transmural lesion formation. This may be attributed to the fact that the CF of the active catheter was controlled around 10 g and the power was controlled at approximately 35W.

When performing BRFA under this study setting, it is considered more reasonable to use AI for the following reasons. First, if BRFA were conducted with a wider range of CF and power, the results might differ. Second, AI is automatically calculated and does not require logarithmic computation. Third, the predictive formula for transmural lesion formation using AI has higher specificity compared to the one using ln (BRFA duration). To further increase the specificity of transmural lesion formation, adding 50 to the calculated AI value from this equation could achieve 100% specificity for transmural lesion formation.

The third notable point of this study is the potential for electrode temperature monitoring to predict steam-pop occurrence. The factors related with steam-pop occurrence during BRFA were previously reported as impedance drop (9) (14), temperature of endocardial catheter (10), RF time (12) and high power (12) but regarding impedance drop, there are also conflicting reports (15). In unipolar ablation, Leshem et al. reported that temperature-controlled ablation using QDOT can create comparable lesion formation without increasing the incidence of steam-pop compared to power-controlled ablation (16). In their study, no steam-pop occurred when the targeted temperature of QDOT was set to 45°C, while steam-pop did occur when set to 50°C, though it was less frequent compared to power-controlled ablation. Nomura et al. reported that regarding DTA, steam-pop occurred only when the catheter tip of the DTA was applied perpendicularly during ablation with a targeted temperature set at 58°C (17). Al-Sheikhli et al. reported that steam-pop and cardiac tamponade occurred in one patient during one of 10 VT ablations with DTA, using a default target temperature of 60°C and an initial power of 50 W (18).

The results of the study showed that temperature-controlled BRFA using QMODE with a cutoff of 45°C could prevent steam-pops on the active catheter side. Additionally, no steam-pops occurred when the temperature of the opposing DTA remained below 55°C. In BRFA, the area most likely to be heated is directly beneath the contact points between the tissue and each of the two electrodes where the radiofrequency current converges, as suggested by computer simulation models. (19) Therefore, monitoring the surface temperature of both electrodes to prevent steam-pop at each location is valuable. From these results, it is suggested that during QMODE ablation set at 45°C, visually monitoring the temperature of the opposing DTA and manually reducing the output of QDT when the temperature exceeds 55°C may help prevent steam-pops on both sides. Using a non-irrigated tip catheter as the return catheter allows for the measurement of the myocardial surface temperature on the opposite side; however, if the tip is not irrigated, there is a risk of thrombosis. While an irrigated tip might cool the temperature sensor itself, DTA, utilizing the high thermal conductivity of diamond, enables effective temperature monitoring. Therefore, using DTA as the return catheter is reasonable. In this study, % Impedance drop was also useful for predicting steam-pop. Saito et al. reported that the frequency of steam-pop decreased when BRFA was performed with reduced power according to the %Imp drop. (20) In this study, it was not verified whether controlling the power output according to DTA temperature could prevent steam-pop. However, in terms of steam-pop occurrence, DTA temperature had much higher specificity compared to % Impedance drop. This result is likely because % Impedance drop can be influenced by the temperature increase in all regions through which the radiofrequency current flows, whereas DTA can measure the temperature specifically in the tissue surface where the catheter tip makes contact, which is near the region most susceptible to steam-pop occurrence.

In the validation of Model 3, lesions were hardly formed when only the distal small tip of the DTA was in contact with the tissue during ablation. This was likely because the proximal larger tip of the DTA was floating in the saline, causing the radiofrequency current to bypass the myocardial tissue and flow from the saline to the proximal larger tip, resulting in insufficient heating of the tissue. The distal small tip is 0.6 mm long, and in practice, it is unlikely that only this electrode would make contact with the tissue. However, if, as indicated by the results of this study, there is little temperature rise in the DTA, it may be worth considering adjusting the positioning of the DTA to ensure effective ablation.

### Study Limitations

This study has some limitations. As mentioned earlier, CF and power were controlled to some extent in this study, and altering the range of these parameters could lead to different results. Second, it is unclear whether similar results would be obtained with BRFA using an inter-electrode distance of 28 mm or more.

## Conclusions

AI adjusted for inter-electrode distance proved to be highly significant for achieving transmural lesion formation during BRFA. Furthermore, temperature-controlled ablation using QDT and DTA demonstrated potential for predicting steam-pop during BRFA.

## Data Availability

The datasets generated during the current study are available from the corresponding author upon reasonable request.

---

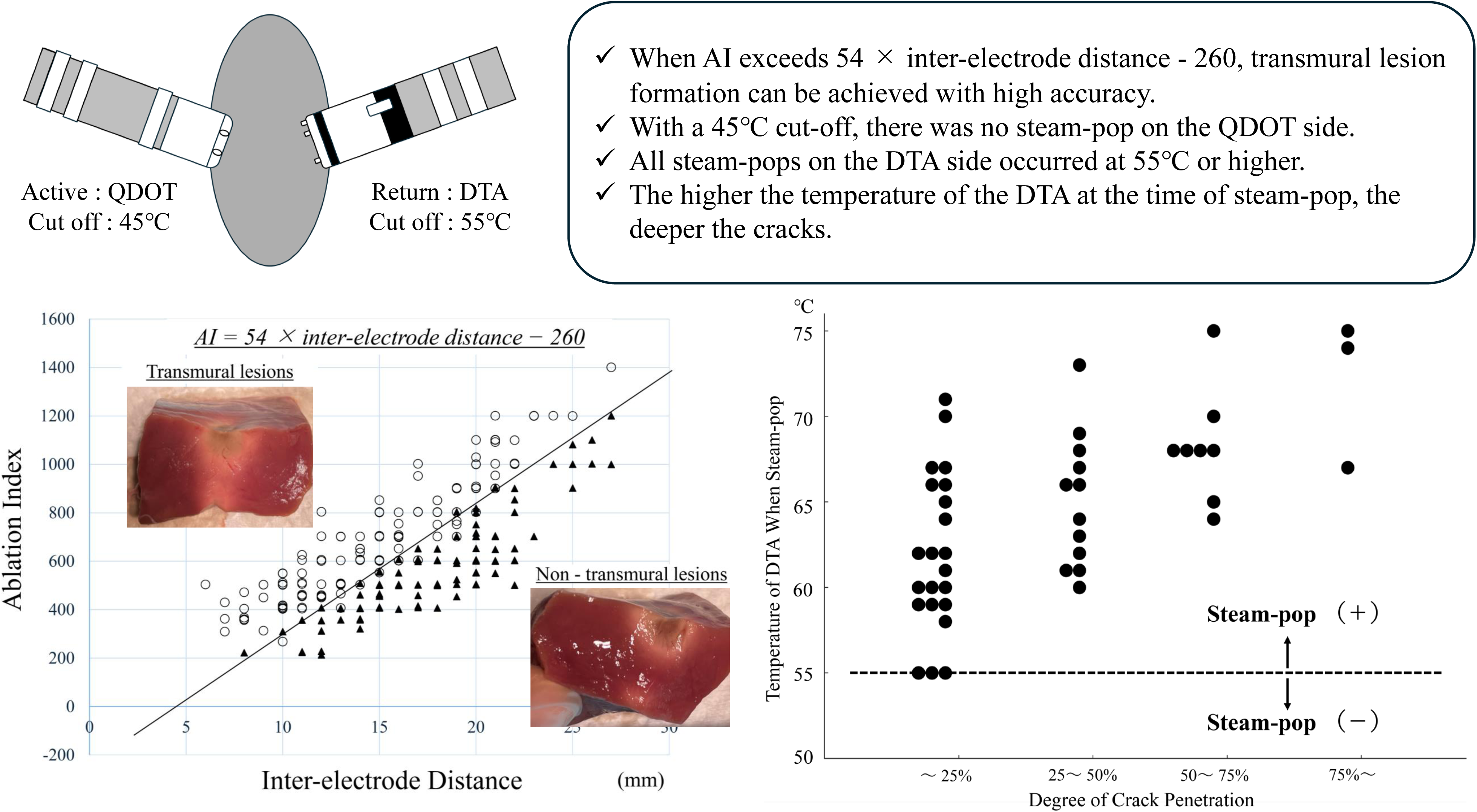

